# DIAGNOSTIC ACCURACY OF ARTIFICIAL INTELLIGENCE FOR ANALYSIS OF 1.3 MILLION MEDICAL IMAGING STUDIES: THE MOSCOW EXPERIMENT ON COMPUTER VISION TECHNOLOGIES

**DOI:** 10.1101/2023.08.31.23294896

**Authors:** Sergey Morozov, Anton Vladzymyrskyy, Natalia Ledikhova, Anna Andreychenko, Kirill Arzamasov, Olga Omelyanskaya, Roman Reshetnikov, Pavel Gelezhe, Ivan Blokhin, Elena Turavilova, Daria Kozhikhina, Daria Anikina, Dmitry Bondarchuk

## Abstract

**Objective:** to assess the diagnostic accuracy of services based on computer vision technologies at the integration and operation stages in Moscow’s Unified Radiological Information Service (URIS).

**Methods:** this is a multicenter diagnostic study of artificial intelligence (AI) services with retrospective and prospective stages. The minimum acceptable criteria levels for the index test were established, justifying the intended clinical application of the investigated index test. The Experiment was based on the infrastructure of the URIS and United Medical Information and Analytical System (UMIAS) of Moscow. Basic functional and diagnostic requirements for the artificial intelligence services and methods for monitoring technological and diagnostic quality were developed. Diagnostic accuracy metrics were calculated and compared.

**Results:** based on the results of the retrospective study, we can conclude that AI services have good result reproducibility on local test sets. The highest and at the same time most balanced metrics were obtained for AI services processing CT scans. All AI services demonstrated a pronounced decrease in diagnostic accuracy in the prospective study. The results indicated a need for further refinement of AI services with additional training on the Moscow population datasets.

**Conclusions:** the diagnostic accuracy and reproducibility of AI services on the reference data are sufficient, however, they are insufficient on the data in routine clinical practice. The AI services that participated in the experiment require a technological improvement, additional training on Moscow population datasets, technical and clinical trials to get a status of a medical device.

## Introduction

From a global perspective, there is an intensive development of artificial intelligence (AI) technologies. AI is associated with the opportunities to improve the quality and availability of medical care, increase productivity and optimize resources. The most developed AI technologies, particularly computer vision, are in the field of diagnostic radiology^1,2^. At the same time, expectations that are unsupported by scientific results may turn out to be superfluous. Levels of investment in the development of new technologies are not a criterion for evidence-based medicine.

A major scientific study is necessitated by a need for a qualitative transformation of the healthcare system, technological and methodological readiness, and demand from the professional community as well as the ongoing hype.

The Russian Federation (RF) has adopted the National Strategy for the Development of Artificial Intelligence for the period until 2030, which contains provisions on the healthcare sector. The application of automated clinical decision support systems is recommended by the regulations of the Ministry of Health of the Russian Federation^3,4^. In 2019, the Moscow Government decided to conduct a large-scale scientific study – “The Experiment on the use of innovative computer vision technologies for the medical image analysis and subsequent application in the Moscow healthcare system” (www.mosmed.ai)^5^.

The Experiment involved legal entities that had developed or had the rights to provide services based on computer vision technologies for the analysis of medical images (AI services). The Experiment was carried out by the Moscow Research and Practical Clinical Center for Diagnostics and Telemedicine Technologies of the Moscow Healthcare Department. The technological component of the Experiment was provided by the Moscow Department of Information Technologies. To support and motivate legal entities representing AI services, the Moscow Government has provided research grants.

Companies from Russia, as well as 65.0% of specialized international companies, were invited to participate in the Experiment. As a result, 21 companies took part in the Experiment, including 8 Russian legal representatives of foreign developers. Under the Experiment terms, each legal entity could submit several separate services for participation. Therefore, the total number of participating AI services was 39 with market coverage in Russia of more than 90%.

We carried out a comprehensive study of diagnostic accuracy and advisability of AI technology applications in radiology as part of the Experiment. It included the survey for radiologists as well as a comparative report turnaround time analysis. This article presents a component of the Experiment dedicated to assessing diagnostic accuracy.

### Study objective

To assess the diagnostic accuracy of services based on computer vision technologies at the integration and operation stages in Moscow’s Unified Radiological Information Service (URIS).

## Material & Methods

STARD-2015 methodology^6^ was used for study reporting.

“The Experiment on the use of innovative computer vision technologies for the medical image analysis and subsequent application in the Moscow healthcare system” was conducted in public outpatient and inpatient healthcare facilities providing primary and specialized medical care for an adult population from January 1, 2020, to December 31, 2020.

### Design

a multicenter diagnostic study of AI services with retrospective and prospective stages^6,7^.

### Inclusion criteria

1. Age over 18 years.
2. Gender male or female.
3. Referral for one of the radiology examination types with results available in the patient medical records.
4. Results of the following imaging studies in the DICOM standard:
  – chest computed tomography (CT) and low-dose computed tomography (LDCT) to detect lung cancer (LC);
  – chest computed tomography to detect COVID-19;
  – chest X-ray (X-ray) and fluorography (FLG) to detect various lung pathology (primarily tuberculosis and lung cancer);
  – chest X-ray to detect COVID-19;
  – mammography (MMG) to detect breast cancer (BC).

### Exclusion criteria

1. A different type of imaging (including a different modality or anatomical area);
2. Absence of signed informed consent for participation in the Experiment.

### Hypothesis

following the method of Korevaar et al.^8^, the minimum acceptable criteria levels for the index test of AI were established, justifying the intended clinical application of the investigated index test:

– sensitivity – 0,81 (with 95% CI);
– specificity – 0,81 (with 95% CI);
– area under the ROC Curve (AUC) – 0,81 (with 95% CI).

***Null hypothesis*** for each study stage – H_0_: {sensitivity D<0,81 and/or specificity <0,81 and/or AUC <0,81}.

### Infrastructure and intervention

The Experiment was based on the infrastructure of the Unified Radiological Information Service and United Medical Information and Analytical System of Moscow (URIS UMIAS). URIS UMIAS is a single digital space, uniting workstations of technicians and radiologists, as well as diagnostic equipment, accumulating information about each study conducted on connected devices^9^. 1,290 diagnostic units were connected to the URIS UMIAS as of January 1, 2020. This corresponds to 100% of digital diagnostic devices of Moscow outpatient and inpatient medical facilities. The opportunity to utilize AI was provided for all radiologists and X-ray technicians, access to study results was ensured for referring physicians. By integrating with the Moscow public services portal (mos.ru), patients were able to get study results (reports and images) through their accounts. By the end of 2020, URIS UMIAS has accumulated more than 7 million reports: computed tomography (CT), magnetic resonance imaging (MRI), radiography (X-ray), fluorography (FLG), mammography (MMG), as well as hybrid studies.

We developed basic functional and diagnostic requirements for the AI services, methods for monitoring technological and diagnostic quality for the Experiment. More than 100 labeled datasets have been generated based on the original methodology. Details of this work will be published elsewhere.

After submitting and approving an application for participation in the Experiment, AI services were integrated into URIS UMIAS. We performed special step-by-step admitting procedures to assess the quality of this process.

After a step-by-step user training, radiologists could have used AI services during routine study reporting.

When a patient sought medical care at public healthcare facilities, a standard examination was carried out, during which an attending physician determined the presence of indications for a radiologic exam and formed a study referral. The study was conducted following the established procedure, and the results were automatically saved in the URIS UMIAS. For the types of studies included in the Experiment, automatic routing to AI service was configured. From a diagnostic device, the studies were routed to only one AI service in a certain period and were automatically processed. The DICOM SR was sent to the URIS UMIAS and was available at the radiologist’s workstation as an additional series. All images in this series were labeled “For research purposes only”. AI services calculated a probability of a target pathology. The final decision in study reporting was always made by a radiologist. When reporting in URIS UMIAS, the radiologist decided whether to open the additional series generated by AI and use the clinical decision support system. A decision to check the additional series was purely voluntary.

### Study stages

1. Retrospective study:
  – index test: AI service integrated into the URIS UMIAS test circuit (n=18);
  – reference test: local test sets. A preparation and local test set tagging was carried out according to the original methodology^10,11^. Sample size was 1,525 studies, grouped into 13 datasets for two types of testing: CT for detecting COVID-19 (n = 200), CT for detecting and classifying COVID-19 with the “CT0-4” grades (n = 325), CT for detecting lung cancer (n = 200), MMG for detecting breast cancer (n = 200), X-ray for detecting various lung pathology (n = 200), X-ray for detecting COVID-19 (n = 200), FLG for detecting COVID-19 (n = 200). The normal-to-pathology ratio in all datasets was 50/50. CT dataset for detecting and classifying COVID-19 with the “CT0-4” grades was an exception with balance of CT-0 : CT-1 : CT-2 : CT-3 : CT-4 equal to 20:20: 20: 20: 20.
2. Prospective study:
  – index test: AI service integrated into the URIS UMIAS, which successfully passed the admitting procedures for processing real clinical data (n=18);
  – reference test: a report of the radiologist who directly conducted a study at the medical facility.

Report analysis was carried out via the in-house developed software “MedLabel” for automated analysis of medical reports^1^. Its primary function was the extraction of keywords denoting signs of pathological changes from textual report data.

Information on sample size for each stage is presented in Table 1.

**Table 1.**
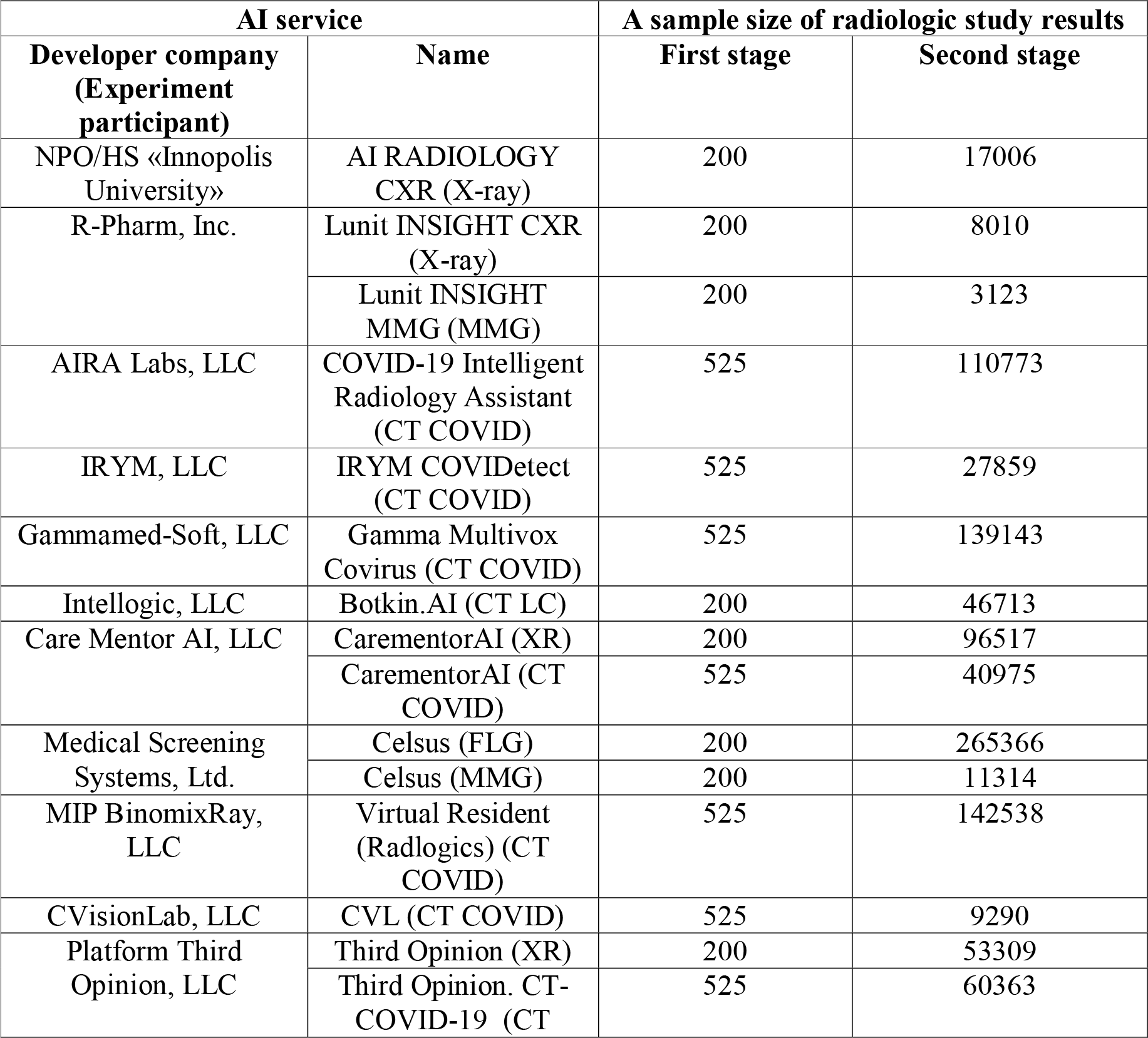
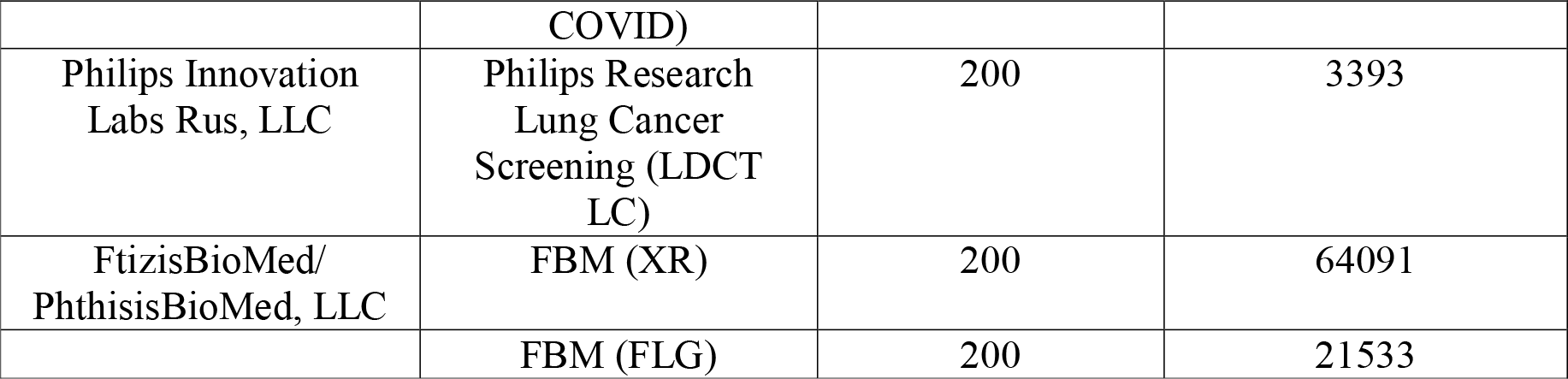
Samples for a step-by-step assessment of diagnostic accuracy of each AI service.

| AI service |  | A sample size of radiologic study results |  |
| --- | --- | --- | --- |
| Developer company<br>(Experiment participant) | Name | First stage | Second stage |
| NPO/HS «Innopolis University» | AI RADIOLOGY CXR (X-ray) | 200 | 17006 |
| R-Pharm, Inc. | Lunit INSIGHT CXR (X-ray) | 200 | 8010 |
|  | Lunit INSIGHT MMG (MMG) | 200 | 3123 |
| AIRA Labs, LLC | COVID-19 Intelligent Radiology Assistant (CT COVID) | 525 | 110773 |
| IRYM, LLC | IRYM COVIDetect (CT COVID) | 525 | 27859 |
| Gammamed-Soft, LLC | Gamma Multivox Covirus (CT COVID) | 525 | 139143 |
| Intellogic, LLC | Botkin.AI (CT LC) | 200 | 46713 |
| Care Mentor AI, LLC | CarementorAI (XR) | 200 | 96517 |
|  | CarementorAI (CT COVID) | 525 | 40975 |
| Medical Screening Systems, Ltd. | Celsus (FLG) | 200 | 265366 |
|  | Celsus (MMG) | 200 | 11314 |
| MIP BinomixRay, LLC | Virtual Resident (Radlogics) (CT COVID) | 525 | 142538 |
| CVisionLab, LLC | CVL (CT COVID) | 525 | 9290 |
| Platform Third Opinion, LLC | Third Opinion (XR) | 200 | 53309 |
|  | Third Opinion. CT-COVID-19 (CT | 525 | 60363 |

<sup>1</sup> Certificate of state registration of the computer program "MedLabel – automated analysis of medical reports" No. 2020664321, application date – 27.10.2020, registration date – 11.11.2020.

|  |  |  |  |
| --- | --- | --- | --- |
|  | COVID) |  |  |
| Philips Innovation Labs Rus, LLC | Philips Research Lung Cancer Screening (LDCT LC) | 200 | 3393 |
| FtizisBioMed/<br>PhthisisBioMed, LLC | FBM (XR) | 200 | 64091 |
|  | FBM (FLG) | 200 | 21533 |

### Statistical analysis

Diagnostic accuracy metrics were calculated and compared^12^, including sensitivity, specificity, area under the ROC curve (AUROC). The threshold was calculated using the Youden index.

### Ethics and registration

The study was approved by the Independent Ethics Committee of the Moscow Society of Radiology on February 20, 2020. The principles for conducting the Experiment corresponded to the Joint Statement of the European and North American Society of Radiologists and others on the Ethics of Artificial Intelligence in Radiology^13^. All patients signed a special informed consent form for voluntary participation. A specially designed brochure was provided for the additional information.

The Experiment was registered in the ClinicalTrials with the assigned ID NCT04489992.

## Results

### Retrospective study

Each AI service integrated into the URIS UMIAS underwent calibration using local test sets. From a scientific point of view, the results enabled us to assess the accuracy of an AI service on real URIS UMIAS data of the Moscow population.

### Prospective study

Each AI service prospectively analyzed incoming studies in the URIS UMIAS. The results were automatically compared with conclusions of radiologists who directly conducted radiologic studies at the medical facilities.

The diagnostic accuracy metrics of AI services are presented in Tables 2, 3, and the Appendix.

**Table 2.** Average diagnostic accuracy metrics for all AI services.

| Parameter | Min | Max | Average value | Median |
| --- | --- | --- | --- | --- |
| **Retrospective study** |  |  |  |  |
| AUC | 0,80 | 0,98 | 0,89±0,06 | 0,91 |
| Sensitivity | 0,68 | 0,94 | 0,85±0,07 | 0,85 |
| Specificity | 0,69 | 0,94 | 0,88±0,07 | 0,89 |
| **Prospective study** |  |  |  |  |
| AUC | 0,63 | 0,85 | 0,74±0,07 | 0,745 |
| Sensitivity | 0,45 | 0,82 | 0,66±0,11 | 0,68 |
| Specificity | 0,62 | 0,93 | 0,76±0,09 | 0,76 |

**Table 3.**
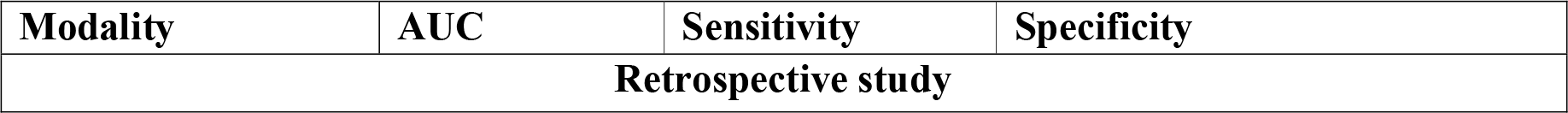
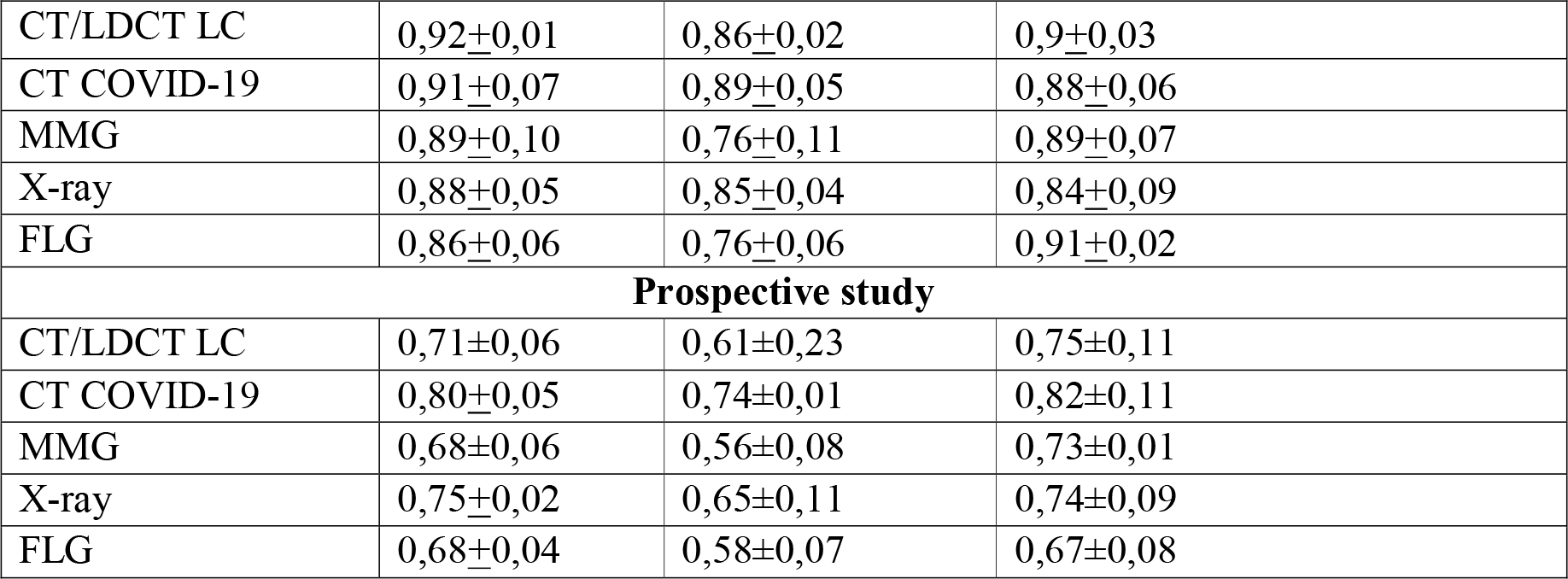
Average diagnostic accuracy metrics by modalities, M+m.

## Discussion

Based on the results of the **retrospective study**, we can conclude that AI Services have good result reproducibility on local test sets. Namely, the deviations of diagnostic metrics did not exceed 10.0% from the initially advertised. A cumulative average of the diagnostic accuracy metrics for all AI services was about 0.9.

It should be noted that AI Services Lunit INSIGHT MMG (MMG), Third OpinionCT-COVID-19 (CT COVID) and COVID-19 Intelligent Radiology Assistant (CT COVID) demonstrated the highest AUC values – 0,96, 0,97 and 0,98, respectively. The lowest AUC values were obtained for FBM (X-ray) – 0,8, Celsus (MMG) – 0,81, and IRYM COVIDetect (CT COVID), CarementorAI (CT COVID) and Celsus (FLG) – 0,82 both.

Modality-wise, the highest and at the same time most balanced metrics were obtained for AI services processing CT scans. Accounting for the specifics of mass chest fluorography, AI services demonstrated low AUC and sensitivity values, and the highest specificity values, which deserve positive recognition.

Diagnostic accuracy in the prospective study has the highest scientific importance, as it allows to evaluate the real-life performance of AI. The circumstances in 2020 included the COVID-19 pandemic resulting in a temporary suspension of screening programs. This affected the types of studies sent to AI services for analysis. Nevertheless, the primary data we obtained were quite sufficient for statistical analysis.

In real clinical conditions, all AI services demonstrated a pronounced decrease in diagnostic accuracy. Only 27.8% (5) of AI services retained a sufficient level of reproducibility (AUC> 0.81 per the requirements for the Experiment). These were the following AI services: COVID-19 Intelligent Radiology Assistant (CT COVID), Lunit INSIGHT CXR (X-ray), Virtual Resident (Radlogics) (CT COVID), Third Opinion CT-COVID-19 (CT COVID), CVL (CT COVID). Most of the AI services – 44.4% (8) had their prospective AUC within 0.7–0.8. The rest fell below 0,69.

Modality-wise, the most stable and relatively high metrics were again shown by AI services analyzing CT scans and radiography. On the other hand, AI services for mammography and fluorography approached critically low values.

A comparison of our results in the Moscow Experiment with prior studies is of interest. Unfortunately, relevant scientific publications have not been found for all AI services; nevertheless, interesting observations have been made.

The Lunit INSIGHT CXR service (X-ray) showed high accuracy and reproducibility in the Experiment, for retrospective and prospective study: AUC 0.91 and 0.82, sensitivity 0.86 and 0.82, specificity 0.85 and 0.70. Our results match the literature data. For patient triage in suspected pulmonary tuberculosis, Lunit INSIGHT CXR (X-ray) demonstrated 88.61% AUC (88.03-89.20). There was a decrease in accuracy for patients over 60 years old or with prior tuberculosis^14^. Slightly worse results were obtained in another study on tuberculosis diagnostics: pooled sensitivity 90.0, specificity 54.1% (44.6-63.3). The sensitivity of the AI service was lower for smear-positive TB cases and HIV infection. The authors concluded that the service can be implemented as a high-sensitivity rule-out test, but users will need threshold scores identified from their patient populations and stratified by HIV- and smear-status^15^. A higher accuracy regarding lung cancer detection on chest X-rays was shown when using Lunit INSIGHT CXR as a clinical decision support system (CDSS)^16,17^. In the retrospective study to identify focal lesions, consolidation, and pneumothorax, the AUC for this AI service was 0.9883, 1.000, and 0.9997, respectively^17^. In one study, Lunit INSIGHT CXR showed fluctuations in AUC, sensitivity, and specificity (0.771-0.872, 69.6-83.3%, 74.0-78.8%, respectively) on a large sample (n = 5887), depending on the type of ground truth. If the results of computed tomography were taken as the ground truth, then the accuracy increased^18^. For the Lunit INSIGHT MMG service, AUC values for retrospective and prospective studies were 0.96 and 0.72. The retrospective study corresponds to the literature data - 0.959 (95% CI 0.952-0.966^19^), but for the prospective study, the AUC was much lower. Such a pronounced decrease has its explanations unrelated to the particular AI service and will be discussed below. The accuracy of radiologists improved when using Lunit INSIGHT MMG as CDSS^19^. Overall, the results for the Lunit INSIGHT service did not contradict the literature data. However, we cannot comment on improving the human reader accuracy via Lunit INSIGHT as CDSS, since it was not a research objective of the Experiment.

The Virtual Resident (Radlogics) service for COVID-19 diagnostics on chest CT demonstrated high accuracy and good result reproducibility. We obtained the following accuracy values for the retrospective and prospective stages: AUC 0.91 and 0.82, sensitivity 0.91 and 0.73, specificity 0.92 and 0.91, respectively. In the prospective stage, the specificity even increased. Prior reported diagnostic accuracy metrics for the Virtual Resident (Radlogics) service were: AUC 0.996 (95% CI 0.989-1.00), sensitivity 98.2%, specificity 92.2%^20^. However, these metrics were established by internal validation on the Chinese population dataset, which was also used to train the algorithm. In the Moscow Experiment, the service had performed without the additional calibration and training. It explains the lower diagnostic values we obtained.

Several services in the Moscow Experiment displayed average results, necessitating a comparison. Botkin.AI service (CT LC) showed AUC levels of 0.91 and 0.66. Only retrospective data was comparable to prior retrospective testing with AUC 0.9274. During the Experiment, sensitivity and specificity were 0.84 and 0.45, 0.92 and 0.83, respectively - significantly lower than claimed by the developers (1.00 and 0.937)^21^. The following data were published for the Celsus service (MMG): the algorithm detected cancer correctly in 45 out of 49 cases (92%)^22^. The absence of generally accepted diagnostic metrics in this publication makes the objective comparison impossible.

We retrospectively tested two services before the Moscow Experiment. Philips Research Lung Cancer Screening (LDCT LC) service demonstrated comparable performance with an initial AUC of 0.930, and at the retrospective stage of the Experiment – 0.92. In the prospective study, AUC decreased to 0.78, followed by specificity (prior – 0.925, retrospectively – 0.87, prospectively – 0.68)^23^. We also previously tested the FBM (FLG) service on two datasets with a balance of normal-to-pathology 50/50 and 95/5 with resulting AUC of 0.74 and 0.64, sensitivity – 0.872 and 0.75, specificity – 0.60 and 0.535, concluding that the accuracy varied depending on the class balance^24^. At the first stage of the Moscow Experiment, the AUC was 0.9 – higher than the prior study. However, due to the limited participation duration of the FBM (FLG) service, it is impossible to conduct a more detailed analysis. It should be noted that an independent group of authors obtained sensitivity – 0.74, specificity – 0.89 in a retrospective study^25^.

We did not find relevant published studies for other AI services.

Overall, the results obtained in the Moscow Experiment indicated a need for further refinement of AI services with additional training on the Moscow population datasets.

A comparison of our results with initial metrics provided by developers is particularly interesting (Table 6). The Experiment is a large-scale independent validation of AI services on new data that were not used to train algorithms. Independent validation, especially in multicenter clinical trials is mandatory for the successful development of artificial intelligence technologies^26^.

**Table 6.** Mean AUC differences (delta) at different stages of AI diagnostic accuracy assessment.

| **AI service** | **Developer-stated minus retrospective** | **Developer-stated minus prospective** | **Retrospective minus prospective** |
| --- | --- | --- | --- |
| AI RADIOLOGY CXR (X-ray) | -0,047 | -0,137 | -0,09 |
| Lunit INSIGHT CXR (X-ray) | -0,01 | -0,1 | -0,09 |
| Lunit INSIGHT MMG (MMG) | 0,06 | -0,18 | -0,24 |
| COVID-19 Intelligent Radiology Assistant (CT COVID) | 0,04 | -0,13 | -0,17 |
| IRYM COVIDetect (CT COVID) | -0,05 | -0,14 | -0,09 |
| Gamma Multivox Covirus (CT COVID) | 0,09 | -0,06 | -0,15 |
| Botkin.AI (CT LC) | -0,03 | -0,28 | -0,25 |
| CarementorAI (BC) | 0,1 | -0,08 | -0,18 |
| CarementorAI (CT COVID) | -0,03 | -0,06 | -0,03 |
| Celsus (FLG) | -0,05 | -0,18 | -0,13 |
| Celsus (MMG) | 0 | -0,18 | -0,18 |
| Virtual Resident (Radlogics) (CT COVID) | 0,1 | 0,01 | -0,09 |
| CVL (CT COVID) | 0,12 | 0,03 | -0,09 |
| Third Opinion (X-ray) | 0,14 | -0,04 | -0,18 |
| Third Opinion. CT-COVID-19 (CT COVID) | 0,02 | -0,13 | -0,15 |
| Philips Research Lung Cancer Screening (LDCT LC) | 0,07 | -0,1 | -0,17 |
| FBM (X-ray) | 0,01 | -0,11 | -0,12 |
| FBM (FLG) | 0,11 | -0,12 | -0,23 |

At the retrospective stage, a slight decrease in AUC of 33.3% (6) AI services was noted in comparison to initially stated metrics. At the same time, most AI services – 66.7% (12) showed a slight increase, and 5 AI services improved their AUC by more than 0,1.

At the prospective stage, only 11.1% (2) of the services maintained AUC values at the initially stated level: Virtual Resident (Radlogics) (CT COVID) and CVL (CT COVID).

Many AI services – 88.9% (16) showed a decrease in AUC, and in 66.7% (12) this deterioration in diagnostic accuracy was 0.1 or more.

Overall, the differences between the AUC values were statistically significant. For both stages, the Mann-Whitney U-test for a given number of compared groups was 99 (p <0.05).

We noted a similar trend when comparing the first and second stages – 100.0% of AI services showed a decrease in the AUC. Less than 0.1 decrease was observed in 33.3% (6) of AI services: CarementorAI (CT COVID), AI RADIOLOGY CXR (X-ray), Lunit INSIGHT CXR (X-ray), IRYM Covidetect (CT COVID), Virtual Resident (Radlogics) (CT COVID), and CVL (CT COVID). On the other hand, FBM (FLG), Lunit INSIGHT MMG (MMG), and Botkin.AI (CT LC) showed a decrease higher than > 0.2. The differences were statistically significant, Mann-Whitney U-test = 99, p <0.05.

A decrease in accuracy can be due to several reasons:

1. Reporting and radiologist’ performance:
  i. AI service detects a pathology that has no clinical significance. Technically, the AI service identifies a finding correctly, however, a doctor does not include it in the report due to low prognostic significance. It leads to an erroneous assessment of the algorithm’s performance as a false-positive.
  ii. A radiologist is guided not only by the image but also by the patient’s clinical data from the electronic medical record. Therefore, the report may differ from a straightforward radiologic image description.
  iii. A radiologist may report a presence of “residual changes” after a disease. Such a report was regarded as containing a pathology, while the AI service no longer detects changes in the image. This leads to an erroneous assessment of the algorithm result as a false-negative.
2. Features of AI service performance:
  i. A low quality of segmentation, leading to a false-positives outside the anatomical target area or even outside the patient’s body, incorrect projection selection (for example, on chest X-ray, a service mistakenly analyzes a lateral projection instead of a frontal one). This is an objective reason for a large number of false-positive results.
  ii. Non-representative datasets used by developers to train algorithms (no studies with technical defects; disease imaging signs are not fully represented). For example, in routine practice, services analyze images after a surgical intervention incorrectly (CXR post lung resection is regarded as a pathology). This is also an objective reason for many false-positive results. Therefore, we may arrive at the following methodological conclusions:
    1. AI in diagnostic radiology should always be verified by a human reader (radiologist).
    2. In the future, with a large-scale implementation and routine application of AI technologies, it is necessary for medical (radiological) information systems to automatically send study results for the independent quality control (audit) regarding the presence of discrepancies between a human reader and AI.

We compared our data on the diagnostic accuracy of AI services by modalities with the published research.

The accuracy of AI services for initial mammography reading is generally comparable to the literature data. In limited samples of several hundred patients, the AI achieved the following results: sensitivity – 86.0-91.0%, specificity – 67.0-79.0%, AUC – 0.89-0.97^27–29^. We recorded a decrease in accuracy at the second stage of the study. However, unlike other authors, we obtained data on the screening mammography AI reading in a prospective multicenter study. Additional training of AI services on the Moscow population datasets is required, especially since fluctuations in the accuracy on datasets of different populations were previously demonstrated^19^. In 2021, Freeman et al published a systematic review on the diagnostic accuracy of AI in breast cancer screening. The review included 12 studies (a total of 131,822 women who underwent mammography), authors did not find any well-designed prospective studies, and the overall methodological quality of included articles was rated as low. Based on the three included articles (79,910 women, 1,878 cancer cases, 36 AI algorithms), 94% of AI algorithms were less accurate than a radiologist; 100% were less accurate than a consensus of two or more human readers. For five included articles (1,086 women, 520 cancer cases, 5 AI algorithms), 100% of AI algorithms are more accurate than a radiologist^30^. Several authors propose augmenting human readers with AI as a clinical decision support system. Therefore, AI may be used for double reading screening mammography^27,28^. The data of our prospective study supports this point of view, noting the need for additional training of AI services and workflow in radiology departments.

The accuracy values of AI services for analyzing CT/LDCT and detecting malignant lesions match the literature data: sensitivity – 73.0-100.0%, specificity – 71.0-89.0%, AUC – 0.86-97.6^31–36^. In a retrospective study on a limited sample, a consistency of AI technologies and medical experts in diagnosing lung nodules with LDCT was established with Cohen’s kappa of 0.846^31^. The retrospective stage of our study was close to the literature data; the prospective stage demonstrated AUCs at the lower limit or even below, sensitivity decreased to 0.61 ± 0.23. The cited authors conducted only retrospective studies on limited datasets, comparing the accuracy of algorithms and a small group of radiologists (up to 10 people). In addition, detection of nodules less than 5 mm by AI may be favorably interpreted. However, such nodules are forwarded to the next round of screening, not requiring any further workup, thus raising doubts about a study methodology. In most cases, studies focused on determining the superior reader of a particular dataset – a human or AI. The purpose of our study was to assess the accuracy and reliability of algorithms in a prospective clinical process, using AI as a technology to support decision-making by a human reader.

In terms of sample size, a retrospective assessment of 5,485 CT scans performed as a part of the National Lung Screening Trial (NLST) in 2002–2004 approaches our findings. For lung nodule detection, the sensitivity of AI was 86.2%, and specificity – 85.0% with higher accuracy of AI in comparison with radiologists^37^. Our results for the prospective stage significantly differ from the literature data. Our study is prospective within the framework of standard processes of radiology departments, while the literature data were obtained retrospectively under conditions of limited testing on small datasets. At the same time, we agree with the conclusions of some authors about the potential applicability of AI technologies for double reading the results of screening studies^36,37^. It is also optimal to use AI as a decision support system. In this context, our research is supported by the work of Zhang et al, 2021, which showed a higher accuracy of AI-assisted reading compared to the human reader reporting only. The accuracy for solid lung nodule detection with AI-assisted reading was 99.1% (versus 86.2%)^38^.

In the pandemic, a significant number of articles and preprints have been published on AI-based COVID-19 detection in chest CT. Many authors use multicenter datasets to develop AI algorithms, as well as test them on new data. Public domain datasets are often used for this purpose^39–42^. In some studies and meta-analyses, high levels of sensitivity 84.0-100.0%, specificity 93.0-100.0%, and AUC 0.92-1.0 were reported^39,41,43–45^. Again, our results at the first stage are fully comparable with the published research – performed via retrospective testing on limited data. However, there is no information on testing algorithms in real clinical settings. Even recent articles published in 2021 on the clinical applicability of AI for detecting COVID-19 on chest CT are based on retrospective multicenter validation using limited-volume datasets^46,47^.

As the pandemic continued, we were creating requirements for AI services on the fly. It quickly became clear that the primary task for AI was segmentation to assist a radiologist in determining disease severity. This decision was also confirmed by an independent publication, indicating the effective AI application for determining the volume of lung parenchymal lesions and forming a radiology report template^48^. However, in our work, we have further advanced this methodology. In the Experiment, AI services should have provided CT COVID-19 analysis results according to the regional classification “CT0-4”^49^, allowing for instant decisions on patient routing.

From a global perspective, the development of automated X-ray analysis for detecting tuberculosis has been underway for over 30 years^50^. Therefore, a significant amount of data has been accumulated, including prospective clinical trials. This is a significant difference between radiography and other modalities.

Some state that AI can assist radiologists in the CXR analysis, and be used as a decision support system for clinicians if human reading is unavailable^51^.

In clinical studies, it is already possible to assess not only AI accuracy in general but also concerning specific patient groups: for example, specificity of an automatic analysis decreases in men with a history of tuberculosis, with increasing age and decreasing body mass index^52^. In the Experiment, as shown above, we observed a decrease in AI diagnostic accuracy between retrospective and prospective stages. In the available studies on the automated analysis of CXR for tuberculosis detection, a difference in diagnostic accuracy was noted. In a systematic review of 53 articles, mean AUC for studies describing algorithms’ development is 0.88 (95% CI 0.82-0.90), while in clinical trials, this indicator decreases to 0.75 (95% CI 0.66-0.87; p = 0.004)^53^. We obtained a similar results at the first retrospective stage, the AUC of AI services was 0.88 + 0.05 (for fluorography – 0.86 + 0.06), and at the second stage, it was 0.75 + 0.02 (0.68 +0.04). A retrospective comparative analysis of the three most accessible AI algorithms for detecting tuberculosis on CXR was performed on a sample of 1,196 patients with the immunological analysis as a reference method. The AUC was 0.92 to 0.94 and the authors recommended including automated analysis in tuberculosis screening programs, especially in limited human resources^14^. It should be noted that one of the algorithms included in the study also took part in the Moscow Experiment. Although the study was multicenter, its limitation was a retrospective nature. Nevertheless, we agree with the authors’ conclusion about the applicability of AI in TB screening. However, we have to emphasize that the suitability of a particular algorithm should be established in the prospective clinical trial.

Overall, testing results of 18 AI services in the prospective study confirmed the previously stated thesis that adjustment or recalibration of AI for the target institution or population is recommended^51^.

Thus, based on the results of the two-stage assessment of AI, the null hypothesis H_0_: {sensitivity <0.81 and/or specificity <0.81 and /or AUC <0.81} in:

– calibration testing on reference datasets was rejected,
– processing CT studies for COVID-19 detection was rejected, for the rest –accepted.

### Limitations

Study limitations are as follows:

1. Datasets for the first stage used an artificial balance of classes, decreasing the significance and clinical relevance of obtained results.
2. Design of the second stage with single radiologist’ report as ground truth, unsupported by expert consensus or peer-review; there were no mechanisms for analyzing reports’ content to exclude biases and defects.
3. Prospective participation duration of AI services in the Experiment was different, affecting a comparative analysis.

## Conclusions

1. The diagnostic accuracy and reproducibility of AI services on the reference data are sufficient, however, they are insufficient on the data in routine clinical practice.
2. All AI serviced demonstrated a pronounced statistically significant (Mann-Whitney U-test = 99, p <0.05) decrease in the diagnostic accuracy below the retrospective values when applied in real clinical conditions. This decline is partially due to methodological problems in the application of AI technologies in clinical practice, as well as some biases.
3. More stable and relatively high values were shown by AI services for computed tomography and radiography; the rest approached critically low values.
4. The null hypothesis H_0_: {sensitivity <0.81 and/or specificity <0.81 and/or AUC <0.81} for calibration testing was rejected; for CT in COVID-19 detection was rejected, for the rest - accepted.
5. The AI services that participated in the Experiment require a technological improvement, additional training on Moscow population datasets, technical and clinical trials to get a status of a medical device.

## Supporting information

title page

## Data Availability

All data produced in the present study are available upon reasonable request to the authors

## Appendix

**Table A1.** Diagnostic accuracy metrics of AI services established at the first assessment stage.

| AI service | AUC |  | Sensitivity |  | Specificity |  |
| --- | --- | --- | --- | --- | --- | --- |
|  | I stage | II stage | I stage | II stage | I stage | II stage |
| AI RADIOLOGY CXR (X-ray)* | 0,74 | 0,77 | 0,4 | 0,56 | 0,96 | 0,83 |
|  | 0,86 | - | 0,83 | - | 0,88 | - |
| Lunit INSIGHT CXR (X-ray) | 0,91 | 0,82 | 0,86 | 0,82 | 0,85 | 0,70 |
| Lunit INSIGHT MMG (MMG) | 0,96 | 0,72 | 0,84 | 0,62 | 0,94 | 0,72 |
| COVID-19 Intelligent Radiology Assistant (CT COVID) | 0,98 | 0,81 | 0,94 | 0,71 | 0,94 | 0,86 |
| IRYM COVIDetect (CT COVID) | 0,82 | 0,73 | 0,84 | 0,72 | 0,78 | 0,71 |
| Gamma Multivox Covirus (CT COVID) | 0,9 | 0,75 | 0,88 | 0,66 | 0,84 | 0,77 |
| Botkin.AI (CT LC) | 0,91 | 0,66 | 0,84 | 0,45 | 0,92 | 0,83 |
| CarementorAI (X-ray) | 0,91 | 0,73 | 0,81 | 0,57 | 0,92 | 0,81 |
| CarementorAI (CT COVID) | 0,82 | 0,79 | 0,82 | 0,82 | 0,82 | 0,76 |
| Celsus (FLG) | 0,82 | 0,69 | 0,72 | 0,53 | 0,9 | 0,73 |
| Celsus (MMG) | 0,81 | 0,63 | 0,68 | 0,50 | 0,84 | 0,74 |
| Virtual Resident (Radlogics) (CT COVID) | 0,91 | 0,82 | 0,91 | 0,73 | 0,92 | 0,91 |
| CVL (CT COVID) | 0,94 | 0,85 | 0,94 | 0,75 | 0,9 | 0,93 |
| Third Opinion (X-ray)* | 0,6 | 0,74 | 0,98 | 0,60 | 0,23 | 0,76 |
|  | 0,92 | - | 0,86 | - | 0,86 | - |
| Third Opinion. CT-COVID-19 (CT COVID)* | 0,86 | 0,82 | 0,78 | 0,78 | 0,86 | 0,80 |
|  | 0,97 | - | 0,9 | - | 0,94 | - |
| Philips Research Lung Cancer Screening (LDCT LC) | 0,92 | 0,75 | 0,87 | 0,78 | 0,88 | 0,68 |
| FBM (X-ray) | 0,8 | 0,68 | 0,9 | 0,70 | 0,69 | 0,62 |
| FBM (FLG)* | 0,77 | 0,67 | 0,74 | 0,63 | 0,76 | 0,62 |
|  | 0,9 | - | 0,8 | - | 0,92 | - |
| * Results of primary and secondary testings are presented. |  |  |  |  |  |  |

## Ethical approval

The study was approved by the Independent Ethics Committee of the Moscow Society of Radiology on February 20, 2020.

## Guarantor

Sergey Morozov MD, PhD, MPH

## Contributorship

SM, AV, NL, AA, and KA conceptualized the foundational principles for this work. A protocol was developed by SM, AV and NL to guide the methodology. AA performed the grey and academic literature searches and KA was the second reviewer for the academic rapid review. All authors were involved in data analysis procedures. PG and IB were responsible for the writing of this manuscript with significant input and review by SM, AV, and KA. All authors agree regarding the final content of the manuscript.

## Acknowledgements

Coordinating, scientific, methodological, and educational tasks of the Experiment were carried out by the staff of the Research and Practical Clinical Center for Diagnostics and Telemedicine Technologies of the Moscow Healthcare Department. As part of the Experiment, a selection of AI & CV services according to established criteria and their integration into the URIS UMIAS were carried out. Technological tasks in the Experiment were solved with the active participation and support of the Moscow Department of Information Technologies.

## Footnotes

1 Certificate of state registration of the computer program “MedLabel – automated analysis of medical reports” No. 2020664321, application date – 27.10.2020, registration date – 11.11.2020.

## REFERENCES

1. Pesapane F, Codari M, Sardanelli F. Artificial intelligence in medical imaging: threat or opportunity? Radiologists again at the forefront of innovation in medicine. Eur Radiol Exp; 2. Epub ahead of print 1 December 2018. doi: 10.1186/S41747-018-0061-6.

2. Ranschaert ER, Morozov S, Algra PR. Artificial intelligence in medical imaging: Opportunities, applications and risks. Artif Intell Med Imaging Oppor Appl Risks 2019; 1–373.

3. Decree of the President of the Russian Federation No. 490 of October 10, 2019 “On the Development of Artificial Intelligence in the Russian Federation”. 2019.

4. Order of the Ministry of Health of the Russian Federation No. 560n of June 9, 2020 ‘On approval of the Regulations for conducting X-ray examinations’. 2020.

5. Regulation of the Government of Moscow No. 1543-PP of November 21, 2019 “On Conducting the Experiment on the Use of Innovative Technologies in the Field of Computer Vision for the Analysis of Medical Images and Subsequent Application in the Healthcare Sys. 2019.

6. Bossuyt PM, Reitsma JB, Bruns DE, et al. STARD 2015: An Updated List of Essential Items for Reporting Diagnostic Accuracy Studies. Clin Chem 2015; 61: 1446–1452.

7. Liu X, Rivera SC, Moher D, et al. Reporting guidelines for clinical trial reports for interventions involving artificial intelligence: the CONSORT-AI Extension. BMJ 2020; 370: m3164.

8. Korevaar DA, Gopalakrishna G, Cohen JF, et al. Targeted test evaluation: a framework for designing diagnostic accuracy studies with clear study hypotheses. Diagnostic Progn Res 2019 31 2019; 3: 1–10.

9. N.S. Polishchuk, N.N. Vetsheva, S.P. Kosarin, S.P. Morozov ESK. Unified Radiological Information Service as a tool for organizational and methodological work of the Research and Practical Clinical Center for Diagnostics and Telemedicine Technologies of the Moscow Healthcare Department (analytical report). Radiol – Pract 2018; 0: 6–17.

10. Morozov SP, Andreychenko AE, Pavlov NA, et al. MosMedData: Chest CT Scans With COVID-19 Related Findings Dataset. Epub ahead of print 13 May 2020. doi: 10.1101/2020.05.20.20100362.

11. Kulberg NS, Gusev MA, Reshetnikov R V., et al. Methodology and tools for creating training samples for artificial intelligence systems for recognizing lung cancer on CT images. Heal care Russ Fed 2020; 64: 343–350.

12. P. Morozov [et. Clinical trials of intelligent technology-based software (diagnostic radiology). In: The best practices of radiology and instrumental diagnostics. 2019, pp. 57–61.

13. Geis JR, Brady A, Wu CC, et al. Ethics of artificial intelligence in radiology: summary of the joint European and North American multisociety statement. Insights Imaging 2019; 10: 1–6.

14. Ahmed MHE S, Sarker MS, Paul MPH K, et al. Tuberculosis detection from chest x-rays for triaging in a high tuberculosis-burden setting: an evaluation of five artificial intelligence algorithms. Lancet Digit Heal 2021; 3: e543–e554.

15. Tavaziva G, Harris M, Abidi SK, et al. Chest X-ray analysis with deep learning-based software as a triage test for pulmonary tuberculosis: an individual patient data meta-analysis of diagnostic accuracy. Clin Infect Dis. Epub ahead of print 21 July 2021. doi: 10.1093/CID/CIAB639.

16. Garcia-Velloso MJ, Ribelles MJ, Rodriguez M, et al. MRI fused with prone FDG PET/CT improves the primary tumour staging of patients with breast cancer. Eur Radiol 2017; 27: 3190–3198.

17. Choi SY, Park S, Kim M, et al. Evaluation of a deep learning-based computer-aided detection algorithm on chest radiographs: Case–control study. Medicine (Baltimore) 2021; 100: e25663.

18. Kim EY, Kim YJ, Choi WJ, et al. Performance of a deep-learning algorithm for referable thoracic abnormalities on chest radiographs: A multicenter study of a health screening cohort. PLoS One 2021; 16: e0246472.

19. Kim HE, Kim HH, Han BK, et al. Changes in cancer detection and false-positive recall in mammography using artificial intelligence: a retrospective, multireader study. Lancet Digit Heal 2020; 2: e138–e148.

20. Gozes O, Ayan Frid-Adar M’, Greenspan H, et al. Rapid AI Development Cycle for the Coronavirus (COVID-19) Pandemic: Initial Results for Automated Detection & Patient Monitoring using Deep Learning CT Image Analysis, https://arxiv.org/abs/2003.05037v3 (2020, accessed 21 November 2021).

21. I.S. Drokin, E.V. Ericheva, O.L. Bukhvalov, P.S. Pilyus, T.S. Malygina VES. Experience in developing and implementing a search system for recognizing malignant tumors using artificial intelligence on the example of computed tomography of the lung. Physicians IT 2019; 3: 48–57.

22. O.E. Karpov, O.Yu. Bronov, A.A. Kapninsky, P.I. Pavlovich, Yu.A. Abovich, S.A. Subbotin, S.V. Sokolova, N.I. Rychagova, A.V. Milova, E.D. NikitinO.E. Karpov, O.Yu. Bronov, A.A. Kapninsky, P.I. Pavlovich, Yu.A. Abovich, S.A. Subbotin, S.V. Sokolova, N.I. R Edn. Comparative study of data analysis results of artificial intelligence based digital mammography system ‘Celsus’ and radiologists. Bull Pirogov Natl Med Surg Cent 2021; 16: 86–92.

23. S.P. Morozov, A.V. Vladzimirsky, V.A. Gombolevsky VGK. Artificial intelligence in lung cancer screening: assessment of a diagnostic accuracy of the algorithm for analyzing low-dose computed tomography. Tuberc Lung Dis 2020; 98: 24–31.

24. Morozov SP, Vladzimirskiy A V., Ledikhova N V., et al. EVALUATION OF DIAGNOSTIC ACCURACY OF THE SYSTEM FOR PULMONARY TUBERCULOSIS SCREENING BASED ON ARTIFICIAL NEURAL NETWORKS. Tuberc Lung Dis 2018; 96: 42–49.

25. P.V. Gavrilov, O.P. Gavrilova UAS. Detection of peripheral lung formations using a program for automated analysis of fluorographic images. Diagnostic Radiol Ther 2020; 1: 77.

26. Baldwin DR, Gustafson J, Pickup L, et al. External validation of a convolutional neural network artificial intelligence tool to predict malignancy in pulmonary nodules. Thorax 2020; 75: 306–312.

27. McKinney SM, Sieniek M, Godbole V, et al. International evaluation of an AI system for breast cancer screening. Nature 2020; 577: 89–94.

28. Rodríguez-Ruiz A, Krupinski E, Mordang JJ, et al. Detection of Breast Cancer with Mammography: Effect of an Artificial Intelligence Support System. Radiology 2019; 290: 305–314.

29. Sasaki M, Tozaki M, Rodríguez-Ruiz A, et al. Artificial intelligence for breast cancer detection in mammography: experience of use of the ScreenPoint Medical Transpara system in 310 Japanese women. Breast Cancer 2020; 27: 642–651.

30. Freeman K, Geppert J, Stinton C, et al. Use of artificial intelligence for image analysis in breast cancer screening programmes: systematic review of test accuracy. BMJ; 374. Epub ahead of print 2 September 2021. doi: 10.1136/BMJ.N1872.

31. Chamberlin J, Kocher MR, Waltz J, et al. Automated detection of lung nodules and coronary artery calcium using artificial intelligence on low-dose CT scans for lung cancer screening: accuracy and prognostic value. BMC Med; 19. Epub ahead of print 1 December 2021. doi: 10.1186/S12916-021-01928-3.

32. Cui S, Ming S, Lin Y, et al. Development and clinical application of deep learning model for lung nodules screening on CT images. Sci Rep; 10. Epub ahead of print 1 December 2020. doi: 10.1038/S41598-020-70629-3.

33. Li X, Guo F, Zhou Z, et al. [Performance of Deep-learning-based Artificial Intelligence on Detection of Pulmonary Nodules in Chest CT]. Zhongguo Fei Ai Za Zhi 2019; 22: 336–340.

34. Li L, Liu Z, Huang H, et al. Evaluating the performance of a deep learning-based computer-aided diagnosis (DL-CAD) system for detecting and characterizing lung nodules: Comparison with the performance of double reading by radiologists. Thorac cancer 2019; 10: 183–192.

35. Liu J kui, Jiang H yang, Gao M di, et al. An Assisted Diagnosis System for Detection of Early Pulmonary Nodule in Computed Tomography Images. J Med Syst; 41. Epub ahead of print 1 February 2017. doi: 10.1007/S10916-016-0669-0.

36. Liang CH, Liu YC, Wu MT, et al. Identifying pulmonary nodules or masses on chest radiography using deep learning: external validation and strategies to improve clinical practice. Clin Radiol 2020; 75: 38–45.

37. Yoo H, Kim KH, Singh R, et al. Validation of a Deep Learning Algorithm for the Detection of Malignant Pulmonary Nodules in Chest Radiographs. JAMA Netw Open 2020; 3: e2017135–e2017135.

38. Zhang Y, Jiang B, Zhang L, et al. Lung Nodule Detectability of Artificial Intelligence-assisted CT Image Reading in Lung Cancer Screening. Curr Med imaging; 17. Epub ahead of print 9 August 2021. doi: 10.2174/1573405617666210806125953.

39. Harmon SA, Sanford TH, Xu S, et al. Artificial intelligence for the detection of COVID-19 pneumonia on chest CT using multinational datasets. Nat Commun; 11. Epub ahead of print 1 December 2020. doi: 10.1038/S41467-020-17971-2.

40. Jin C, Chen W, Cao Y, et al. Development and evaluation of an artificial intelligence system for COVID-19 diagnosis. Nat Commun 2020 111 2020; 11: 1–14.

41. Li L, Qin L, Xu Z, et al. Using Artificial Intelligence to Detect COVID-19 and Community-acquired Pneumonia Based on Pulmonary CT: Evaluation of the Diagnostic Accuracy. Radiology 2020; 296: E65–E71.

42. Sushentsev N, Bura V, Kotnik M, et al. A head-to-head comparison of the intra- and interobserver agreement of COVID-RADS and CO-RADS grading systems in a population with high estimated prevalence of COVID-19. BJR open 2020; 2: 20200053.

43. Lessmann N, Sánchez CI, Beenen L, et al. Automated assessment of COVID-19 reporting and data system and chest CT severity scores in patients suspected of having COVID-19 using artificial intelligence. Radiology 2021; 298: E18–E28.

44. Mei X, Lee HC, Diao K yue, et al. Artificial intelligence-enabled rapid diagnosis of patients with COVID-19. Nat Med 2020; 26: 1224–1228.

45. Ozsahin I, Sekeroglu B, Musa MS, et al. Review on Diagnosis of COVID-19 from Chest CT Images Using Artificial Intelligence. Comput Math Methods Med; 2020. Epub ahead of print 2020. doi: 10.1155/2020/9756518.

46. Ardakani AA, Kwee RM, Mirza-Aghazadeh-Attari M, et al. A practical artificial intelligence system to diagnose COVID-19 using computed tomography: A multinational external validation study. Pattern Recognit Lett 2021; 152: 42–49.

47. Zhang F. Application of machine learning in CT images and X-rays of COVID-19 pneumonia. Medicine (Baltimore) 2021; 100: e26855.

48. Belfiore MP, Urraro F, Grassi R, et al. Artificial intelligence to codify lung CT in Covid-19 patients. Radiol Med 2020; 125: 500–504.

49. Morozov SP, Andreychenko AE, Pavlov NA, et al. MosMedData: Chest CT Scans with COVID-19 Related Findings Dataset. medRxiv 2020; 2020.05.20.20100362.

50. Kulkarni S, Jha S. Artificial Intelligence, Radiology, and Tuberculosis: A Review. Acad Radiol 2020; 27: 71–75.

51. Hwang EJ, Goo JM, Yoon SH, et al. Use of Artificial Intelligence-Based Software as Medical Devices for Chest Radiography: A Position Paper from the Korean Society of Thoracic Radiology. Korean J Radiol; 22. Epub ahead of print 13 September 2021. doi: 10.3348/KJR.2021.0544.

52. Khan FA, Majidulla A, Tavaziva G, et al. Chest x-ray analysis with deep learning-based software as a triage test for pulmonary tuberculosis: a prospective study of diagnostic accuracy for culture-confirmed disease. Lancet Digit Heal 2020; 2: e573–e581.

53. Harris M, Qi A, Jeagal L, et al. A systematic review of the diagnostic accuracy of artificial intelligence-based computer programs to analyze chest x-rays for pulmonary tuberculosis. PLoS One; 14. Epub ahead of print 1 September 2019. doi: 10.1371/JOURNAL.PONE.0221339.

