## Supplementary material for "DIAGNOSTIC ACCURACY OF ARTIFICIAL INTELLIGENCE FOR ANALYSIS OF 1.3 MILLION MEDICAL IMAGING STUDIES: THE MOSCOW EXPERIMENT ON COMPUTER VISION TECHNOLOGIES": title page

### ^1^ State Budget-Funded Health Care Institution of the City of Moscow «Research and Practical Clinical Center for Diagnostics and Telemedicine Technologies of the Moscow Health Care Department»

Sergey Morozov, head of Research and Practical Clinical Center for Diagnostics and Telemedicine Technologies of the Moscow Health Care, ORCID 0000-0001-6545-6170,

Anton Vladzymyrskyy, deputy director of Research and Practical Clinical Center for Diagnostics and Telemedicine Technologies of the Moscow Health Care, ORCID 0000-0002-2990-7736,

Natalia Ledikhova, deputy director of Research and Practical Clinical Center for Diagnostics and Telemedicine Technologies of the Moscow Health Care, ORCID 0000-0002-1446-424X,

Anna Andreychenko, deputy director of Research and Practical Clinical Center for Diagnostics and Telemedicine Technologies of the Moscow Health Care, ORCID 0000-0001-6359-0763,

Kirill Arzamasov, senior research assistant of Research and Practical Clinical Center for Diagnostics and Telemedicine Technologies of the Moscow Health Care, ORCID 0000-0001-7786-0349,

Olga Omelyanskaya, deputy director of Research and Practical Clinical Center for Diagnostics and Telemedicine Technologies of the Moscow Health Care, ORCID 0000-0002-0245-4431,

Roman Reshetnikov, deputy director of Research and Practical Clinical Center for Diagnostics and Telemedicine Technologies of the Moscow Health Care, ORCID 0000-0002-9661-0254,

Pavel Gelezhe, research assistant of Research and Practical Clinical Center for Diagnostics and Telemedicine Technologies of the Moscow Health Care, ORCID 0000-0003-1072-2202,

Ivan Blokhin, research assistant of Research and Practical Clinical Center for Diagnostics and Telemedicine Technologies of the Moscow Health Care, ORCID 0000-0002-2681-9378,

Elena Turavilova, deputy director of Research and Practical Clinical Center for Diagnostics and Telemedicine Technologies of the Moscow Health Care, ORCID 0000-0002-2403-8332,

Daria Kozhikhina, radiologist of Research and Practical Clinical Center for Diagnostics and Telemedicine Technologies of the Moscow Health Care, ORCID 0000-0001-7690-8427,

Daria Anikina, radiologist of Research and Practical Clinical Center for Diagnostics and Telemedicine Technologies of the Moscow Health Care, ORCID 0000-0001-6554-4779,

Dmitry Bondarchuk, radiologist of Research and Practical Clinical Center for Diagnostics and Telemedicine Technologies of the Moscow Health Care, ORCID 0000-0001-8752-0591,
